# Age-specific findings on lifestyle and trajectories of cognitive function from the Korean Longitudinal Study of Aging

**DOI:** 10.1101/2023.03.26.23287772

**Authors:** Seungju Lim, Eunyoung Yoo, Ickpyo Hong, Jihyuk Park

## Abstract

**Background:** There is a paucity of longitudinal studies in the literature which have investigated age-related differences in the relationship between lifestyle factors and cognitive decline. Thus, the purpose of this study was to investigate which lifestyle factors at baseline would slow the longitudinal rate of cognitive decline in young-old (55-64 years), middle-old (65-74 years), and old-old (75+ years) individuals.

**Methods:** We conducted an 11-year follow-up of 6189 older adults from the Korean Longitudinal Study of Aging, which is a cohort of community-dwelling older Koreans. Lifestyle factors including physical activity (PA), social activity (SA), smoking, and alcohol consumption were assessed at baseline. Cognitive function was measured at 2-year intervals over a duration of 11 years. Latent growth curve modeling (LGM) and multigroup analyses (MGA) were performed.

**Results:** The influence of lifestyle factors on the rate of cognitive decline differed by age. Smoking at baseline (−0.05, 95% CI −0.11 to −0.001, per study wave) accelerated the cognitive decline in young-old individuals while frequent SA participation at baseline (0.02, 95% CI 0.01 to 0.03, per study wave) decelerated the cognitive decline in middle-old individuals. None of the lifestyle factors in this study decelerated the cognitive decline in old-old individuals.

**Conclusion:** Cognitive strategies through modifiable lifestyle factors such as smoking cessation in the young-old and frequent SA participation in the middle-old age groups may have great potential in preventing cognitive decline. Since the influence of lifestyle factors varies by age group, age-specific approaches are recommended for promoting cognitive health.

**What is already known on this topic:** Previous studies have shown that lifestyle factors are associated with cognitive decline in older adults. However, longitudinal population-based studies are scarce, and few studies have considered age differences based on the heterogeneity of the elderly population.

**What this study adds:** The results of the present study show a difference in the influence of lifestyle factors on the rate of cognitive decline with age. Smoking at baseline accelerated cognitive decline in the young-old (55-64 years) group, while frequent social activity participation at baseline decelerated in the middle-old (65-74 years) group. In the old-old group (75+ years), there were no lifestyle factors that could decelerate cognitive decline.

**How this study might affect research, practice or policy:** These results emphasize the importance of an age-specific modifiable lifestyle approach for promoting cognitive health in the elderly.

## INTRODUCTION

Decline in cognitive function is a major issue associated with aging [1]. Studies have been focusing on identifying modifiable lifestyle factors that could preserve cognitive function and prevent cognitive decline in the elderly population [2]. Lifestyle refers to an individual’s multifaceted health habits [4] including physical activity (PA) [3], social activity (SA) [4], smoking [5], and drinking habits [6]. Elderly community dwellers can maintain cognitive function during the aging process by adopting an everyday healthy lifestyle. According to “use it or still lose it” hypothesis, unhealthy lifestyle including non-exercising, social disengagement, smoking and drinking, were related to declined cognitive functioning in comparison to healthy lifestyle habits such as exercising, social engagement, non-smoking and non-drinking [5,6]. However, evidence derived from cross-sectional studies may be limited to draw conclusions regarding a more complex association of change [2]. Therefore, longitudinal studies are required to determine the relationship between lifestyle factors and age-related cognitive decline.

Recent longitudinal studies have reported the impact of lifestyle on cognition. PA at baseline has been associated with the rate of change in cognitive function, and it was reported that physically inactive older adults experienced a greater decline in cognitive function compared to the reference group [7,8]. Meta-analysis of longitudinal studies demonstrated a favorable relationship between PA and cognitive trajectories during the follow-up period [9,10]. However, the impact of lifestyle factors, such as social engagement, smoking, and alcohol consumption, on cognitive decline has not been consistently reported. Studies have reported that SA at baseline may help delay cognitive decline [11,12] whereas other studies have failed to prove so [13,14]. Similarly, a reciprocal relationship study [15] reported that cognitive function could modify subsequent changes in confiding and practical social support, but not vice versa. Other lifestyle studies have demonstrated that smoking [16] and consuming alcohol [16] had significant effect on accelerating the rates of decline in cognitive function, whereas another study showed that global cognition was not associated with consuming alcohol at any point over long periods [17]. Even some well-designed randomized controlled trials failed to find significant effects of PA on cognitive function [18–20]. These mixed findings of longitudinal studies may be attributed to the heterogeneity of the elderly population.

The elderly population is not a biologically and socially homogeneous group [21]. The rate of cognitive decline has been reported to accelerate with the stage of ageing, which means that the decline slope is steeper in the oldest-old group than in the young-old age group [22]. The young-old group engaged more in SA than the oldest-old group because they were usually working and not retired yet [23]. This multifaceted heterogeneity may influence the effects of lifestyle factors on cognitive decline. The target lifestyle factors for prevention of dementia have been reported to change according to the lifespan. Education is a modifiable risk factor for dementia in early life, whereas smoking and social isolation are modifiable risk factors in late life [24]. Some differences have been reported between the young-old and old-old groups in terms of the risk factors for memory decline [24]. Fewer self-maintenance activities and lesser SA were found to be the lifestyle risk factors in the young-old and old-old age groups, respectively. These studies suggest that lifestyle factors that prevent cognitive decline are diverse and depend on the stage of aging [24]. Despite evidence of the association between lifestyle factors and cognitive function, the present evidence is insufficient to determine which lifestyle factors decelerate cognitive decline in different stages of aging, such as young-old, middle-old, and old-old age groups. It may be challenging to identify the key lifestyle factors that affect changes in cognitive decline at different stages of aging. Latent growth modeling is a powerful tool for analyzing the relationship between lifestyle factors and changes in cognitive decline. Therefore, the purpose of this study was to investigate the lifestyle factors that slowed the rate of cognitive decline in different age groups.

## METHODS

### Study sample and procedures

The data for this study were obtained from the 2^nd^ (2008) to 7^th^ wave (2018) of the Korean Longitudinal Study of Ageing (KLoSA). This survey is a nationally representative longitudinal study of community-dwelling older adults. Since 2006, the Korea Labor Institute has been collecting data on 10,254 community-dwelling older adults aged ≥ 45 years from all the regions around Korea except Jeju island every 2 years, with the average sample retention rate of 77.1% until 2018 [25]. The KLoSA collected information regarding health status, PA, SA, and health behaviors. For the present study, we extracted data of approximately 11 years, from 2008 to 2018, from the KLoSA database. The exclusion criteria were: age < 55 years at baseline, missing values of cognitive function at any time point, and missing values of lifestyle factors at baseline. Upon follow-up, the analysis of this study included 6,189 participants in 2008, 85.94% in 2010, 93.38% in 2012, 92.45% in 2014, 93.03% in 2016, and 89.47% in 2018. The study participants were classified into three groups on the basis of the baseline age: young-old (55-65y, N=2,346); middle-old (55-65y, N=2,407); and old-old (75y or more, N=1,436). Informed consent was obtained from all the participants before commencement of the study. All the procedures in this study were conducted in accordance with the tenets of Declaration of Helsinki.

### Cognitive function

Cognitive function of the study participants over 11 years (2008 to 2018) was measured using the Korean version of the Mini-Mental State Examination (K-MMSE) score [26]. The K-MMSE is a validated measure of cognitive function in the Korean population, based on the original version of the MMSE [27]. The K-MMSE measures time/spatial orientation, memory registration, attention and calculation, memory recall, language, and visual construction functions. It includes a total of 19 questions with 30 points. Higher scores indicate better cognitive function. In this study, the Cronbach’s alpha reliability was estimated to be approximately 0.799.

### Lifestyle factors

Lifestyle factors in this study included PA, SA, smoking, and alcohol consumption. All lifestyle variables used in this study were measured at baseline (2008). PA at baseline was measured by asking the respondents about their **regular exercise status** and **regular exercise duration**. Regular exercise status was assessed by asking the respondents whether they exercised regularly at least once a week. Responses were dichotomized into 1 (regular exercise) and 0 (no-exercise) [2]. Regular exercise duration was assessed by asking the respondents how long they exercised. The responses were categorized as 0 (none), 1 (<1 year), 2 (1–2 years), 3 (3–4 years), 4 (5-6 years), 5 (≥ 7 years) [28]. SA at baseline was measured by enquiring about the respondents’ **diversity and frequency of SA participation**. The diversity of SA participation was assessed by asking whether they participated in the following six types of SA: (1) Religious activities; (2) Friendship activities; (3) Leisure, Culture, or Sports Clubs; (4) Family or School Reunion; (5) Volunteer works; and (6) Political activities. The responses were summed and categorized into 0 (no participation), 1 (one activity), and 2 (≥ two activities) [29]. The frequency of SA participation was assessed by asking the participants how often they participated in the above activities. Higher scores indicated higher frequency of SA participation. **Alcohol consumption** responses were dichotomized into 1 (current drinker) and 0 (never-drinker) [2]. **Smoking status** responses were also dichotomized into 1 (current smoker) and 0 (never-smoker) [2]. For the six observed variables, responses such as “not returned,” “no answer,” “N/A,” and “don’t know”, were considered missing values and removed.

### Covariates

All covariates used in this study were measured at the baseline (2008). Age, sex, educational level, marital status, residence, log-transformed household income, and employment status were measured. **Self-rated health** was measured using a 5□point Likert scale wherein the scores ranged from 1 (very good) to 5 (very bad). Higher scores were reverse coded to indicate higher self-rated health [2]. **Depressive symptoms** were measured using an adaptation of the 10-item Center for Epidemiologic Studies Depression Scale (CES-D) [30]. A 4□point Likert scale from 0 (rarely or none of the time) to 3 (most of the time) was used, and each response was dichotomized into 0 (rarely or none of the time) and 1 (sometimes, often, most of the time) [31]. The scores were summed, and higher scores indicated more severe depression.

### Statistical analysis

Descriptive statistics of the demographics were used to examine the characteristics of the study participants. Latent growth curve modeling (LGM) and multigroup analysis (MGA) were then performed. The LGM consists of unconditional and conditional models. First, an unconditional model was used to examine the initial level (i.e., intercept) and rate of change (i.e., slope) of cognitive function. To determine the most accurate model for estimating cognitive trajectory, we conducted a chi-square difference test between the no-growth and linear slope models. Second, MGA between young-old, middle-old, and old-old groups was performed to examine the age differences in cognitive trajectories. Third, once the significance of the variance of the intercept and slope of the cognitive trajectories were verified, a conditional model was used to examine the association of the initial level and rates of change in cognitive function with lifestyle factors along with the effect of lifestyle factors on the intercept and slope of cognitive function. Lastly, MGA was performed to examine the impact of age differences on lifestyle factors in cognitive trajectories. The goodness of model fit was determined by comparative fit index (CFI; ≥ 0.90), Tucker and Lewis Index (TLI; ≥ 0.90), root mean square error of approximation (RMSEA; ≤ 0.06), and standardized root mean square residual (SRMR; ≤ 0.08) [32]. Chi-square statistic (χ2) was reported in this manuscript but not used to determine the model fit due to its high sensitivity to large sample size [32]. Missing data were handled using full information maximum likelihood (FIML). IBM SPSS version 25.0 and MPLUS 8.8 were used for statistical analyses.

## RESULTS

### Descriptive results

The sample comprised 2346, 2407, and 1436 participants in the young-old, middle-old, and old-old age groups (Table 1). Significant differences were found in terms of age according to analysis of variance (ANOVA), even after the analysis. Participants in the young-old age group tended to be highly educated, married, wealthier, self-rated themselves as healthier, less likely to be depressed, more likely to exercise regularly, more likely to participate in SA, more likely to smoke, and more likely to consume alcohol.

**Table 1.**
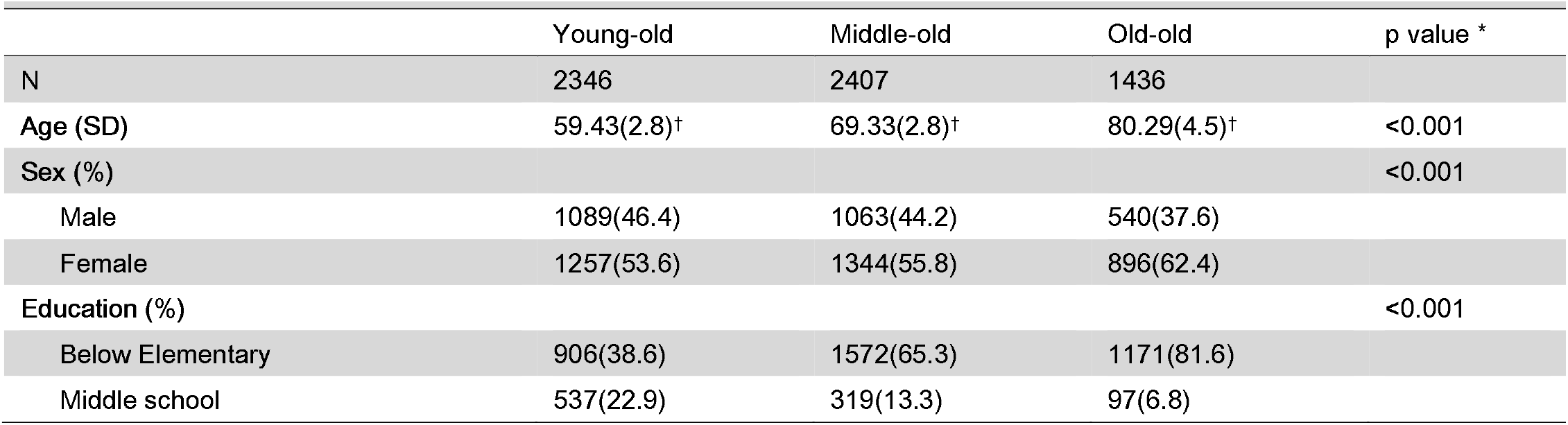
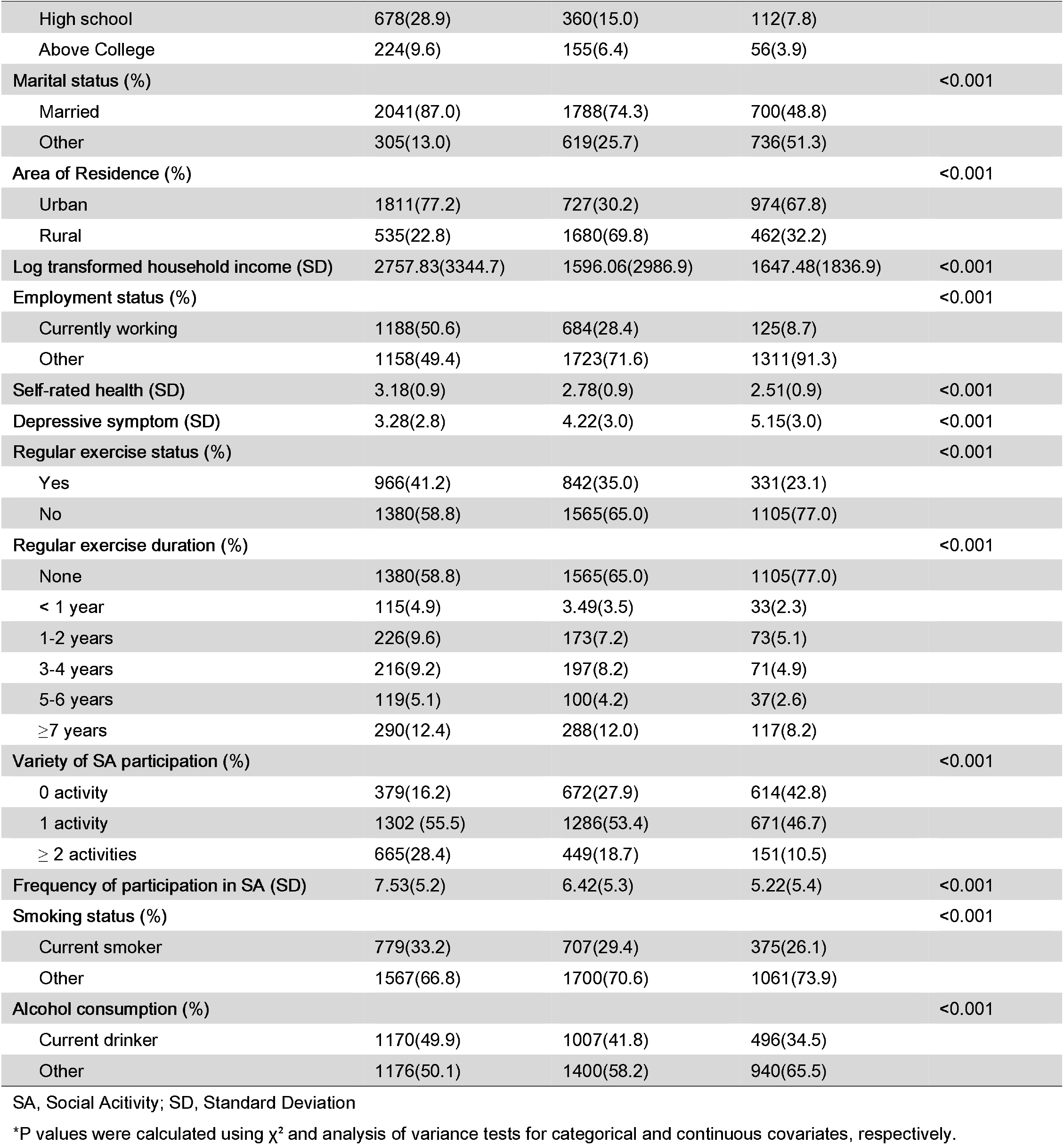
Baseline characteristics of the participants according to the age-group

### Estimation of cognitive trajectories

We performed a chi-square difference test between the linear slope model and the no-growth model to determine the most accurate trajectory shape. Significant difference was found between the linear slope model (χ2 [df] = 274.43 [16]; p < 0.001; CFI = 0.99, TLI = 0.99, SRMR = 0.02, RMSEA = 0.05) and no-growth model (χ2 [df] = 2077.72 [19]; p < 0.001; CFI = 0.89, TLI = 0.89, SRMR = 0.19, RMSEA = 0.13), suggesting that the linear slope models adequately represented the cognitive trajectories.

An unconditional linear slope model of LGM was used to identify the cognitive trajectory of the participants. Their cognitive trajectory began at K□MMSE score of 24.32, and the rate of change was found to be −0.20 for the 2□year interval (χ2 [df]=274.43 [16], p<.001; CFI=0.99; TLI=0.99; SRMR=0.02; RMSEA=0.05). The unconditional LGM with MGA showed significant differences according to the age groups (Δχ2 [df]=356.10 [48], p<.001; CFI=0.98; TLI=0.98; SRMR=0.04; RMSEA=0.06), with the sharpest declining pattern evident in the old-old group with time. The significance of covariance of intercept and slope was observed in the young-old and middle-old groups (Table 2).

**Table 2.** Intercept and slope of cognitive trajectories

|  | Total | Young-old | Middle-old | Old-old |
| --- | --- | --- | --- | --- |
| Intercept mean | 24.32 (24.18 to 24.45) | 26.86 (26.72 to 26.99) | 24.43 (24.24 to 24.63) | 19.97 (19.62 to 20.32) |
| Slope mean | $-0.20$ ($-0.21$ to $-0.18$) | $-0.09$ ($-0.11$ to $-0.07$) | $-0.20$ ($-0.23$ to $-0.17$) | $-0.44$ ($-0.49$ to $-0.38$) |
| Intercept variance | 25.02 (23.91 to 26.13) | 7.89 (7.23 to 8.54) | 18.31 (16.93 to 19.68) | 34.58 (31.11 to 38.06) |
| Slope variance | 0.17 (0.16 to 0.19) | 0.09 (0.08 to 0.10) | 0.19 (0.16 to 0.21) | 0.29 (0.22 to 0.37) |
| Intercept-slope covariance | 0.06 (0.001 to 0.11) | $-0.14$ ($-0.22$ to $-0.06$) | $-0.10$ ($-0.17$ to $-0.02$) | $-0.08$ ($-0.21$ to 0.05) |

\*The estimates are $\beta$ regression coefficient (95% CI).

### Effects of lifestyle factors on the cognitive trajectories

Conditional LGM was performed to identify the effects of lifestyle factors on the intercepts and slopes of cognitive function of the participants. After adjusting the covariates in Model 2 (χ2 [df]=477.15 [76], p<.001; CFI=0.98; TLI=0.98; SRMR=0.01; RMSEA=0.03), participants with longer duration of regular exercise and frequent SA participation at the baseline showed high cognitive intercepts. Frequency of SA participation decreased the cognitive decline by 0.01. Smoking at baseline accelerated the cognitive decline by about −0.05.

The conditional LGM with MGA showed significant differences according to age group after adjusting for covariates (Δχ2 [df]=664.47 [228], p<.001; CFI=0.97; TLI=0.96; SRMR=0.02; RMSEA=0.03). Individuals in the young-old group with longer duration of regular exercise and frequent SA participation at the baseline showed high cognitive intercepts. However, smoking in the young-old group accelerated cognitive decline by approximately −0.05. Individuals in the middle-old group who regularly exercised, frequently participated in SA, and smoked at the baseline showed high cognitive intercepts. The frequency of SA participation was the only factor that decelerated cognitive decline by 0.02. Individuals in the old-old group with longer duration of regular exercise and frequent SA participation at the baseline showed high cognitive intercepts; however, no lifestyle factors decelerated cognitive decline (Table 3).

**Table 3.** Effects of lifestyle factors on intercepts and slopes

|  | Total | Young-old | Middle-old | Old-old |
| --- | --- | --- | --- | --- |
| **Effects on intercepts** |  |  |  |  |
| Regular exercise |  |  |  |  |
| Model 1† | 1.04 (0.47 to 1.61) | -0.32 (-0.85 to 0.20) | 0.85 (0.01 to 1.69) | 0.18 (-1.56 to 1.93) |
| Model 2‡ | 0.20 (-0.29 to 0.68) | -0.29 (-0.78 to 0.20) | 0.95 (0.16 to 1.73) | -0.53 (-2.09 to 1.04) |
| Regular exercise duration |  |  |  |  |
| Model 1† | 0.33 (0.18 to 0.48) | 0.35 (0.20 to 0.49) | 0.33 (0.10 to 0.55) | 0.87 (0.40 to 1.33) |
| Model 2‡ | 0.25 (0.12 to 0.38) | 0.23 (0.10 to 0.37) | 0.10 (-0.11 to 0.31) | 0.44 (0.02 to 0.85) |
| Variety of SA participation |  |  |  |  |
| Model 1† | 1.89 (1.60 to 2.19) | 0.28 (-0.001 to 0.56) | 0.97 (0.54 to 1.41) | 2.51 (1.60 to 3.43) |
| Model 2‡ | -0.01 (-0.26 to 0.25) | -0.14 (-0.4 to 0.13) | 0.15 (-0.26 to 0.57) | 0.65 (-0.19 to 1.50) |
| Frequency of SA participation |  |  |  |  |
| Model 1† | 0.08 (0.03 to 0.14) | 0.17 (0.11 to 0.24) | 0.1 (0.02 to 0.19) | 0.07 (-0.07 to 0.21) |
| Model 2‡ | 0.20 (0.15 to 0.25) | 0.15 (0.09 to 0.20) | 0.13 (0.06 to 0.21) | 0.18 (0.05 to 0.31) |
| Smoking status |  |  |  |  |
| Model 1† | 1.27 (0.96 to 1.59) | 0.42 (0.11 to 0.73) | 1.59 (1.13 to 2.05) | 2.13 (1.31 to 2.95) |
| Model 2‡ | 0.44 (0.14 to 0.75) | 0.19 (-0.15 to 0.54) | 0.996 (0.52 to 1.48) | 0.489 (-0.31 to 1.29) |
| Alcohol consumption |  |  |  |  |
| Model 1† | 0.92 (0.63 to 1.21) | 0.47 (0.18 to 0.77) | 0.51 (0.08 to 0.94) | 0.68 (-0.07 to 1.44) |
| Model 2‡ | -0.06 (-0.33 to 0.2) | 0.25 (-0.04 to 0.54) | -0.22 (-0.66 to 0.21) | -0.45 (-1.181 to 0.28) |
| **Effects on Slopes** |  |  |  |  |
| Regular exercise |  |  |  |  |
| Model 1† | -0.02 (-0.09 to 0.05) | 0.00 (-0.07 to 0.08) | -0.09 (-0.20 to 0.03) | -0.27 (-0.55 to 0.02) |
| Model 2‡ | -0.06 (-0.13 to 0.01) | 0.01 (-0.07 to 0.08) | -0.11 (-0.23 to 0.01) | -0.26 (-0.549 to 0.02) |
| Regular exercise duration |  |  |  |  |
| Model 1† | -0.02 (-0.03 to 0.003) | -0.01 (-0.03 to 0.01) | -0.01 (-0.04 to 0.02) | 0.04 (-0.03 to 0.11) |
| Model 2‡ | -0.01 (-0.03 to 0.01) | -0.01 (-0.03 to 0.01) | -0.01 (-0.04 to 0.02) | 0.05 (-0.101 to 0.19) |
| Variety of SA participation |  |  |  |  |
| Model 1† | 0.06 (0.02 to 0.09) | 0.01 (-0.03 to 0.05) | -0.01 (-0.07 to 0.05) | 0.05 (-0.1 to 0.19) |
| Model 2‡ | 0.00 (-0.04 to 0.04) | 0.00 (-0.04 to 0.04) | -0.02 (-0.079 to 0.04) | 0.05 (-0.101 to 0.20) |
| Frequency of SA participation |  |  |  |  |
| Model 1† | 0.004 (-0.004 to 0.01) | 0.01 (-0.003 to 0.01) | 0.02 (0.01 to 0.03) | -0.01 (-0.03 to 0.01) |
| Model 2‡ | 0.01 (0.001 to 0.02) | 0.01 (0.00 to 0.02) | 0.02 (0.01 to 0.03) | -0.01 (-0.032 to 0.02) |
| Smoking status |  |  |  |  |
| Model 1† | -0.02 (-0.06 to 0.02) | 0 (-0.05 to 0.04) | -0.03 (-0.1 to 0.03) | -0.06 (-0.21 to 0.09) |
| Model 2‡ | -0.05 (-0.1 to -0.01) | -0.05 (-0.11 to -0.001) | -0.035 (-0.11 to 0.04) | -0.10 (-0.26 to 0.06) |
| Alcohol consumption |  |  |  |  |
| Model 1† | 0.02 (-0.02 to 0.05) | -0.02 (-0.06 to 0.03) | -0.01 (-0.07 to 0.05) | 0.05 (-0.09 to 0.18) |
| Model 2‡ | -0.02 (-0.05 to 0.03) | -0.03 (-0.08 to 0.01) | -0.01 (-0.08 to 0.06) | 0.04 (-0.11 to 0.18) |

SA, Social Activity
\*The estimates are $\beta$ regression coefficient (95% CI).
†Model 1 is a crude model
‡Model 2 is adjusted for age, sex, education, marital status, residence, log transformed household income, employment status, self-rated health, and depressive symptoms

## DISCUSSION

The purpose of this study was to examine which lifestyle factors at baseline slowed the rate of cognitive decline in different age groups. Results showed a difference in the influence of lifestyle factors on the rate of cognitive decline with age. Smoking at baseline accelerated cognitive decline in the young-old group, whereas frequent SA participation at baseline delayed the cognitive decline in the middle-old group. None of the lifestyle factors in this study decelerated cognitive decline in the old-old group.

Different lifestyle factors were found to affect the rate of cognitive decline in the young-old and middle-old groups. The results revealed that smoking at baseline was associated with accelerated cognitive decline in the young-old group, whereas frequent participation in SAs at baseline was associated with slower cognitive decline in the middle-old group. This finding was in concordance with previous studies that reported that individuals in Young-old group who smoked at baseline experienced faster cognitive decline in global cognition [33]. In the middle-old group, only the frequency of SA participation, not diversity, decelerated the cognitive decline. A previous longitudinal systematic review and meta-analysis [34] confirmed that frequent social interaction in the middle-old age is important to prevent cognitive decline. Although the effects of diversity and frequency of SA participation on cognitive decline have been widely discussed [35], recent studies have reported that frequency was more sensitive than diversity when predicting cognitive abilities [36]. In particular, individuals in the middle-old group have more time to engage in SA compared to the young-old group since they belong to the post-retirement age [23]. The social and biological heterogeneity of the age groups may have resulted in these different effects of lifestyle factors on cognitive decline. In addition, the old-old group demonstrated different results than the young- and middle-old groups.

We did not find a significant association between lifestyle factors and cognitive decline in the old-old group, which was not in concordance with the findings of previous studies. Even in individuals over the age of 75 years, some predictors of cognitive trajectories associated with slower cognitive decline [37] have been reported as modifiable lifestyle factors (such as, PA, SA, and health behaviors). However, the race and ethnicity of individuals comprising the study population in the previous studies included in the systematic review differed from those in the studies based on the Korean population. Race and ethnicity differences in the research population could explain why lifestyle factors were not associated with cognitive decline in the old-old group. In addition, the KLoSA data used in this study have been regarded as the representative data and reflect the cultural and environmental characteristics of Korean individuals belonging to the old-old group through systematic sampling with probability proportional to size [25]. In addition to race and ethnic differences, the lifestyle measurement method may also have led to different findings from previous research regarding the association between PA and cognitive decline.

In contrast to previous research, we found that the PA level was not associated with decreased rate of cognitive decline throughout the stages of aging. Although a meta-analysis of prospective cohort studies [38,39] found that PA is protective against higher rate of cognitive decline, a mix of the components of PA (i.e., housework, gardening, cleaning the car), limited sample population of sex (male/female), and age (aged 65 years) should also be acknowledged. A few studies have focused on regular exercise, a representative measurement variable of PA, rather than covering overall PA. Since this study focused only on exercise and not on broad PA, the results regarding the influence of PA on cognitive decline could differ from those of previous studies [40]. In addition, we need to consider the limitations of measurement accuracy of overall PA using a questionnaire. Further longitudinal studies beginning at young-old or earlier age group, including both sexes, can help solve the unanswered hypothesis about whether PA is related to longitudinal cognitive decline.

This study is the first to investigate the age-specific effects of lifestyle on longitudinal cognitive decline among young-old, middle-old, and old-old groups in a population-based Korean sample. Although this study was strengthened by a well-powered longitudinal design, it included time-invariant lifestyle factors measured at baseline. To identify more dynamic longitudinal associations between changes in lifestyle factors and cognitive decline, future studies should consider time-varying lifestyle factors. Also, further studies are required to determine the association between specific cognitive domains and lifestyle factors among young-old, middle-old, and old-old groups.

In conclusion, different lifestyle factors delayed the rate of cognitive decline over the 11-year follow-up period in the young-old, middle-old, and old-old groups. Prevention of cognitive decline through modifiable lifestyle factors, such as smoking cessation in the young-old and frequent SA participation in the middle-old groups, may have great potential in preventing cognitive decline. Since the influence of lifestyle factors varies by age group, age-specific approaches are recommended for promoting cognitive health.

## Data Availability

The Korean Longitudinal Study of Aging can be accessed online (https://survey.keis.or.kr/klosa/klosa01.jsp)

## Acknowledgements

We are grateful to all the researchers who participated in this study and the entire Korean Longitudinal Study of Aging team, including the interviewers, technicians, clerical workers, research scientists, volunteers, managers, and receptionists.

## Contributors

SJL, EYY, IPH and JHP conceived and designed the study. SJL wrote the first draft of the manuscript and conducted data analyses. EYY and IPH revised the final manuscript. JHP supervised the analyses and drafted the manuscript. All the authors critically reviewed and approved the final version of this manuscript. JHP is responsible for the overall content as a guarantor.

## Funding

This work was supported by the Ministry of Education of the Republic of Korea and the National Research Foundation of Korea (NRF-2021S1A3A2A02096338)

## Competing interests

None declared

## Patient consent for publication

Not applicable.

## Ethics approval

This study involved human participants and the Korean Longitudinal Study of Aging was approved by the Korea Labor Institute (Approval number: 33602). All the participants provided informed consent to participate before commencement of the study. The authors obtained approval for secondary data analyses from the Yonsei University Institutional Review Board (Approval number: 1041849-202005-SB-067-01).

## Provenance and peer review

Not commissioned; externally peer reviewed

## Notes

### Competing Interest Statement

The authors have declared no competing interest.

### Author Declarations

Institutional Review Board of Yonsei University gave ethical approval for this work Board (Approval number: 1041849-202005-SB-067-01)

## REFERENCES

1 Clouston SAP, Brewster P, Kuh D, et al. The dynamic relationship between physical function and cognition in longitudinal aging cohorts. Epidemiol Rev 2013;35:33–50. doi:10.1093/epirev/mxs004

2 Lim S-J, Park J-H. The Study of the Convergent Factors of the Lifestyle on the Cognitive Decline among Elderly. J Korea Converg Soc 2020;11:229–36. doi:10.15207/JKCS.2020.11.8.229

3 Cohrdes C, Mensink GBM, Hölling H. How you live is how you feel? Positive associations between different lifestyle factors, cognitive functioning, and health-related quality of life across adulthood. Qual Life Res 2018;27:3281–92. doi:10.1007/s11136-018-1971-8

4 Dause T, Kirby E. Aging gracefully: Social engagement joins exercise and enrichment as a key lifestyle factor in resistance to age-related cognitive decline. Neural Regen Res 2019;14:39–42. doi:10.4103/1673-5374.243698

5 Yu J, Collinson SL, Liew TM, et al. Super-cognition in aging: Cognitive profiles and associated lifestyle factors. Appl Neuropsychol 2020;27:497–503. doi:10.1080/23279095.2019.1570928

6 Liu T, Luo H, Tang JYM, et al. Does lifestyle matter? Individual lifestyle factors and their additive effects associated with cognitive function in older men and women. Aging Ment Heal 2020;24:405–12. doi:10.1080/13607863.2018.1539833

7 Hamer M, Terrera GM, Demakakos P. Physical activity and trajectories in cognitive function: English longitudinal study of ageing. J Epidemiol Community Health 2018;72:477–83. doi:10.1136/jech-2017-210228

8 Min JW. A longitudinal study of cognitive trajectories and its factors for koreans aged 60 and over: A latent growth mixture model. Int J Geriatr Psychiatry 2018;33:755–62. doi:10.1002/gps.4855

9 Blondell SJ, Hammersley-mather R, Veerman JL. Does physical activity prevent cognitive decline and dementia?: A systematic review and meta-analysis of longitudinal studies. BMC Public Health 2014;14:1–12. doi:10.1186/1471-2458-14-510

10 Sofi F, Valecchi D, Bacci D, et al. Physical activity and risk of cognitive decline: A meta-analysis of prospective studies. J Intern Med 2011;269:107–17. doi:10.1111/j.1365-2796.2010.02281.x

11 Bae SM. The Mediating Effect of Physical Function Decline on the Association Between Social Activity and Cognitive Function in Middle and Older Korean Adults: Analyzing Ten Years of Data Through Multivariate Latent Growth Modeling. Front Psychol 2020;11:2008. doi:10.3389/fpsyg.2020.02008

12 Zhou Y, Chen Z, Shaw I, et al. Association between social participation and cognitive function among middle- and old-aged Chinese: A fixed-effects analysis. J Glob Health 2020;10:1–11. doi:10.7189/jogh.10.020801

13 Aartsen MJ, Smits CHM, Van Tilburg T, et al. Activity in Older Adults: Cause or Consequence of Cognitive Functioning? A Longitudinal Study on Everyday Activities and Cognitive Performance in Older Adults. J Gerontol B Psychol Sci Soc Sci 2002;57:P153–62. doi:10.1093/geronb/57.2.p153

14 Brown CL, Gibbons LE, Kennison RF, et al. Social activity and cognitive functioning over time: A coordinated analysis of four longitudinal studies. J Aging Res 2012;2012:287438. doi:10.1155/2012/287438

15 Liao J, Muniz-terrera G, Head J, et al. Dynamic Longitudinal Associations Between Social Support and Cognitive Function□: A Prospective Investigation of the Directionality of Associations. J Gerontol B Psychol Sci Soc Sci 2018;73:1233–43. doi:10.1093/geronb/gbw135

16 Hagger-Johnson G, Bell S, Britton A, et al. Cigarette smoking and alcohol drinking in a representative sample of English school pupils: Cross-sectional and longitudinal associations. Prev Med (Baltim) 2013;56:304–8. doi:10.1016/j.ypmed.2013.02.004

17 Gross AL, Rebok GW, Ford DE, et al. Alcohol consumption and domain-specific cognitive function in older adults: Longitudinal data from the Johns Hopkins precursors study. J Gerontol B Psychol Sci Soc Sci 2011;66:39–47. doi:10.1093/geronb/gbq062

18 Sink KM, Espeland MA, Castro CM, et al. Effect of a 24-month physical activity intervention vs health education on cognitive outcomes in sedentary older adults: The LIFE randomized trial. JAMA 2015;314:781–90. doi:10.1001/jama.2015.9617

19 Ngandu T, Lehtisalo J, Solomon A, et al. A 2 year multidomain intervention of diet, exercise, cognitive training, and vascular risk monitoring versus control to prevent cognitive decline in at-risk elderly people (FINGER): A randomised controlled trial. Lancet 2015;385:2255–63. doi:10.1016/S0140-6736(15)60461-5

20 Barnes DE, Santos-Modesitt W, Poelke G, et al. The mental activity and exercise (MAX) trial: A randomized controlled trial to enhance cognitive function in older adults. JAMA Intern Med 2013;173:797–804. doi:10.1001/jamainternmed.2013.189

21 Lowsky DJ, Olshansky SJ, Bhattacharya J, et al. Heterogeneity in healthy aging. J Gerontol A Biol Sci Med Sci 2014;69:640–9. doi:10.1093/gerona/glt162

22 Zhao H, Wen W, Cheng J, et al. An accelerated degeneration of white matter microstructure and networks in the nondemented old–old. Cereb Cortex 2022;bhac372. doi:10.1093/cercor/bhac372

23 Neugarten BL. Age Groups in American Society and the Rise of the Young-Old. Ann Am Acad Pol Soc Sci 1974;415:187–98. doi:10.1177/000271627441500114

24 Livingston G, Huntley J, Sommerlad A, et al. Dementia prevention, intervention, and care: 2020 report of the Lancet Commission. Lancet, 2020;396:413–46. doi:10.1016/S0140-6736(20)30367-6

25 Institute KL. User Guide for 2020 Korean Longitudinal Study of Ageing. Seoul: Korea Labor Institute. 2020.

26 Kang Y, Na DL, Hahn S. A validity study on the Korean Mini-Mental State Examination (K-MMSE) in dementia patients. J Korean Neurol Assoc 1997;15:300–8.

27 Folstein MF, Folstein SE. “Mini-mental state”: a practical method for grading the cognitive state of patients for the clinician. 1975;12:189–98.

28 Kim JH. Regular physical exercise and its association with depression: A population-based study short title: Exercise and depression. Psychiatry Res 2022;309:114406. doi:10.1016/j.psychres.2022.114406

29 Choi Y, Park S, Cho KH, et al. A change in social activity affect cognitive function in middle-aged and older Koreans: analysis of a Korean longitudinal study on aging (2006–2012). Int J Geriatr Psychiatry 2016;31:912–9. doi:10.1002/gps.4408

30 Radolff LS. The CES-D scale: a self-report depression scale for research in the general population. Appl Psychol Meas 1977;1:385–401.

31 Lee H-J. Social activity participation and congnitive function in middle and older adults: A longitudinal study on the reciprocal relationship. Mental Health and Social Work. Ment Heal Soc Work 2015;43:138–67.

32 Hu LT, Bentler PM. Cutoff criteria for fit indexes in covariance structure analysis□: Conventional criteria versus new alternatives. Structural Equation Modeling 1999;6:1–55.

33 Sabia S, Elbaz A, Dugravot A, et al. Impact of smoking on cognitive decline in early old age: The Whitehall II cohort study. Arch Gen Psychiatry 2012;69:627–35. doi:10.1001/archgenpsychiatry.2011.2016

34 Evans IEM, Martyr A, Collins R, et al. Social Isolation and Cognitive Function in Later Life: A Systematic Review and Meta-Analysis. J Alzheimers Dis 2019;70:S119–44. doi:10.3233/JAD-180501

35 Lee S, Charles ST, Almeida DM. Change Is Good for the Brain: Activity Diversity and Cognitive Functioning across Adulthood. J Gerontol B Psychol Sci Soc Sci 2021;76:1036–48. doi:10.1093/geronb/gbaa020

36 Bielak AAM, Mogle JA, Sliwinski MJ. Two sides of the same coin? Association of variety and frequency of activity with cognition. Psychol Aging 2019;34:457–66. doi:10.1037/pag0000350

37 Wu Z, Phyo AZZ, Al-harbi T, et al. Distinct Cognitive Trajectories in Late Life and Associated Predictors and Outcomes: A Systematic Review. J Alzheimers Dis Reports 2020;4:459–78. doi:10.3233/adr-200232

38 Beydoun MA, Beydoun HA, Gamaldo AA et al. Epidemiologic studies of modifiable factors associated with cognition and dementia: systematic review and meta-analysis. BMC Public Health 2014;14:643. https://doi.org/10.1186/1471-2458-14-643

39 Sofi F, Valecchi D, Bacci D, et al. Physical activity and risk of cognitive decline: A meta-analysis of prospective studies. J Intern Med 2011;269:107–17. doi:10.1111/j.1365-2796.2010.02281.x

40 Yaffe K, Barnes D, Nevitt M, et al. A Prospective Study of Physical Activity and Cognitive Decline in Elderly Women: Women Who Walk. Arch Intern Med 2001;161:1703–8. doi:10.1001/archinte.161.14.1703

